# Improved prediction of schizophrenia by leveraging genetic overlap with brain morphology

**DOI:** 10.1101/2020.08.03.20167510

**Authors:** Dennis van der Meer, Alexey A. Shadrin, Kevin O’Connell, Francesco Bettella, Srdjan Djurovic, Thomas Wolfers, Dag Alnæs, Ingrid Agartz, Olav B. Smeland, Ingrid Melle, Jennifer Monereo Sánchez, David E.J. Linden, Anders M. Dale, Lars T. Westlye, Ole A. Andreassen, Oleksandr Frei, Tobias Kaufmann

**Affiliations:** NORMENT, Division of Mental Health and Addiction, Oslo University Hospital & Institute of Clinical Medicine, University of Oslo, Oslo, Norway; School of Mental Health and Neuroscience, Faculty of Health, Medicine and Life Sciences, Maastricht University, Maastricht, The Netherlands; Department of Medical Genetics, Oslo University Hospital, Oslo, Norway; NORMENT, Department of Clinical Science, University of Bergen, Bergen, Norway; Department of Radiology and Nuclear Medicine, Maastricht University Medical Center, Maastricht, The Netherlands; Center for Multimodal Imaging and Genetics, University of California at San Diego, La Jolla, CA 92037, USA; Department of Psychology, University of Oslo, Oslo, Norway; Centre for Bioinformatics, Department of Informatics, University of Oslo, Oslo, Norway

**Keywords:** Genome-wide association study, Brain morphology

## Abstract

Schizophrenia is a complex, polygenic disorder associated with subtle, distributed abnormalities in brain morphology. Here, we report large genetic overlap between schizophrenia and brain morphology, which enabled derivation of polygenic risk scores more predictive of schizophrenia diagnosis than the current state-of-the-art. Our results illustrate the potential of exploiting genetic overlap in imaging genetics studies, and how pleiotropy-enriched risk scores may improve prediction of polygenic brain disorders.

---

Schizophrenia is a highly heritable neurodevelopmental disorder with a complex polygenic architecture, involving numerous common polymorphisms with small effect sizes.^1^ While genome-wide association studies (GWAS) have identified hundreds of genetic variants that are significantly associated with schizophrenia, together these explain only a fraction of its heritability.^2^ Identifying more variants may allow for improved prediction of schizophrenia diagnosis from genetic data, with potential clinical value. However this is currently not within reach, as uncovering a significant portion of the heritability with conventional approaches has been estimated to require sample sizes of more than a million individuals.^3^

The complexity of schizophrenia’s genetic architecture is reflected in its association with brain morphology. At the group level, brain imaging studies have documented thinner cerebral cortex in patients with schizophrenia compared to healthy peers, particularly encompassing frontal and temporal regions, and a mostly global, non-specific, reduction in cortical surface area.^4^ Subcortically, the disorder is associated most notably with smaller hippocampal and amygdala volumes, as well as larger lateral ventricles^5^ and smaller cerebellum.^6^ In line with the clinical and genetic heterogeneity of schizophrenia, imaging studies have documented substantial brain structural heterogeneity,^7^ and specific regional deviations are only shared by a few percent of individuals.^8^

It is possible to boost the power of schizophrenia genetic studies by taking advantage of genetic overlap with related traits,^9^ such as brain morphology. However, large-scale investigations have surprisingly shown no or low genetic correlations between schizophrenia and brain structure.^10,11^ Substantial genetic overlap may remain undetected by measures of genetic correlation due to mixed directions of effects of the shared genetic variants across the two traits cancelling each other out.^12^ Conditional false discovery rate (cFDR) analysis leverages overlapping SNP associations, regardless of direction of effects, to re-rank the test statistics in a primary phenotype conditional on the associations in a secondary phenotype.^9,13^ This approach has been shown to strongly improve GWAS yield.^13^ Further, brain morphology genetics may be analyzed more powerfully by using a multivariate approach, which capitalizes on the shared genetic signal across brain measures by taking into account that the brain is an integrated unit.^14^

Here we investigated whether there is significant genetic overlap between schizophrenia and brain morphology, and whether this overlap can be leveraged to improve polygenic prediction of schizophrenia. For this, we processed T1-weighted magnetic resonance image (MRI) scans from 33,735 individuals with White European ancestry (mean age 64.3, standard deviation (SD) 7.5, 52.0% female), as part of the UK Biobank (UKB),^15,16^ using the standardized pipeline of FreeSurfer.^17,18^ We extracted a total of 175 regional and global brain measures, including surface area, cortical thickness and subcortical volumes, see Supplementary Table 1. For all measures, we regressed out age, sex, scanning site, a proxy of segmentation quality (FreeSurfer’s Euler number),^19^ and the first twenty genetic principal components. Making use of the UKB v3 imputed data,^20^ we performed a GWAS on all the pre-residualized measures through the multivariate omnibus statistical test (MOSTest), yielding one set of summary statistics that captures the significance of its association across all 175 brain morphology measures for each SNP.^14^ We subsequently conditioned the wave 2 summary statistics of the Psychiatric Genomics Consortium GWAS of schizophrenia, (PGC2),^2^ on our brain morphology GWAS results through the cFDR analysis, i.e. a pleiotropy-informed approach.

We identified 571 independent genetic loci significantly associated with brain morphology through our multivariate GWAS approach, see Figure 1a.

**Figure 1.**
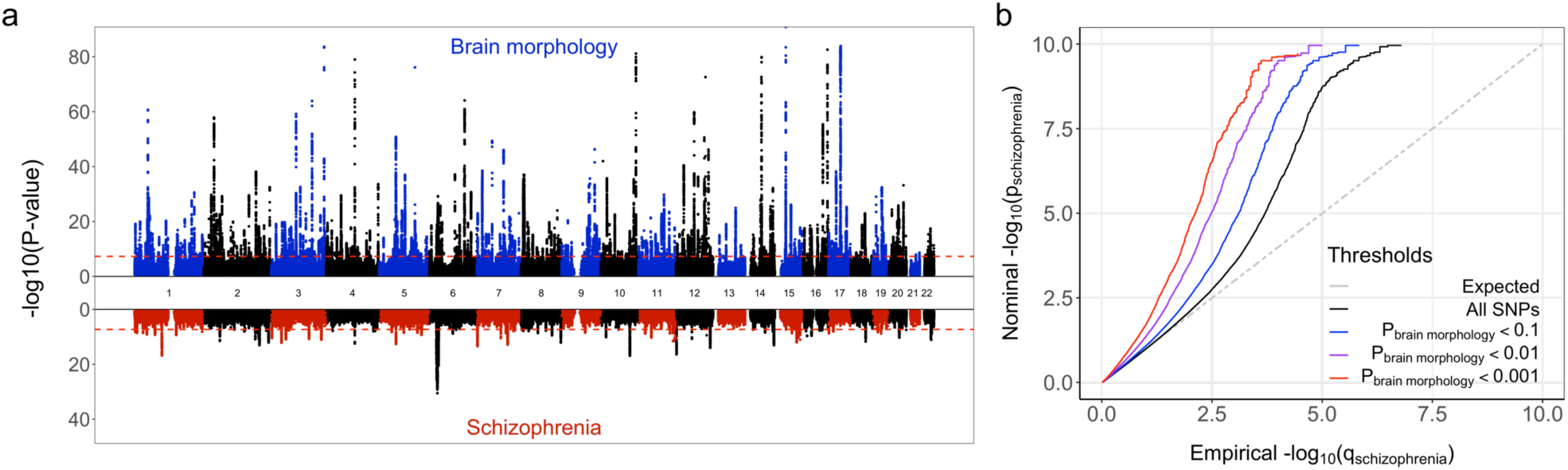
Genetic overlap between schizophrenia and brain morphology. a) Miami plot, depicting the -log10(p-values), on the y-axis, of each genetic variant by their genomic position per chromosome on the x-axis, as identified through the GWAS on brain morphology (top half) and schizophrenia (bottom half). The red dashed lines indicate the genome-wide significance threshold of 5×10^−8^. Note, y-axis is clipped at -log10(p-value)=100. b) Quantile-quantile plot, conditioning the PGC2 schizophrenia GWAS on the multivariate brain morphology GWAS. Successive leftward deflection from the null distribution (dotted line) of the observed p-value distributions at more stringent significance thresholds (color-coded lines) indicates genetic overlap between the two traits.

Of these, 24 loci were also among the 96 loci (i.e. 25%) that were genome-wide significant in the PGC2 schizophrenia GWAS.

Through gene-based analyses using Multi-marker Analysis of GenoMic Annotation (MAGMA),^21,22^ we further found 1952 genes, of which 261 were among the 475 genes (i.e. 55%) identified in the PGC2 schizophrenia data. Supplementary Data 1 and 2 list all discovered loci and genes.

The cFDR analysis of the PGC2 schizophrenia GWAS and the multivariate brain morphology GWAS further confirmed considerable genetic overlap, see Figure 1b. Utilizing this overlap, we identified 295 independent significant loci, of which 201 were not reported in the original PGC2 schizophrenia GWAS, see Supplementary Figure 1 and Supplementary Data 3. Supplementary Figure 2 provides the results of additional analyses, listing the genetic overlap between the individual univariate brain measures and schizophrenia.

The 295 loci identified by the pleiotropy-informed cFDR analysis were mapped onto 836 genes, which were significantly enriched for genes with known upregulated expression in the brain, particularly in the cerebellum and frontal cortex, but not in other tissues. This is contrary to the genes identified through the brain morphology GWAS and the PGC2 schizophrenia GWAS, which showed little specificity of gene expression for any tissue, see Figure 2. The mapped genes were further highly significantly overrepresented in gene sets previously identified through GWAS of bipolar disorder, neuroticism, cognitive ability, and intelligence, see Supplementary Figure 4.

**Figure 2.**
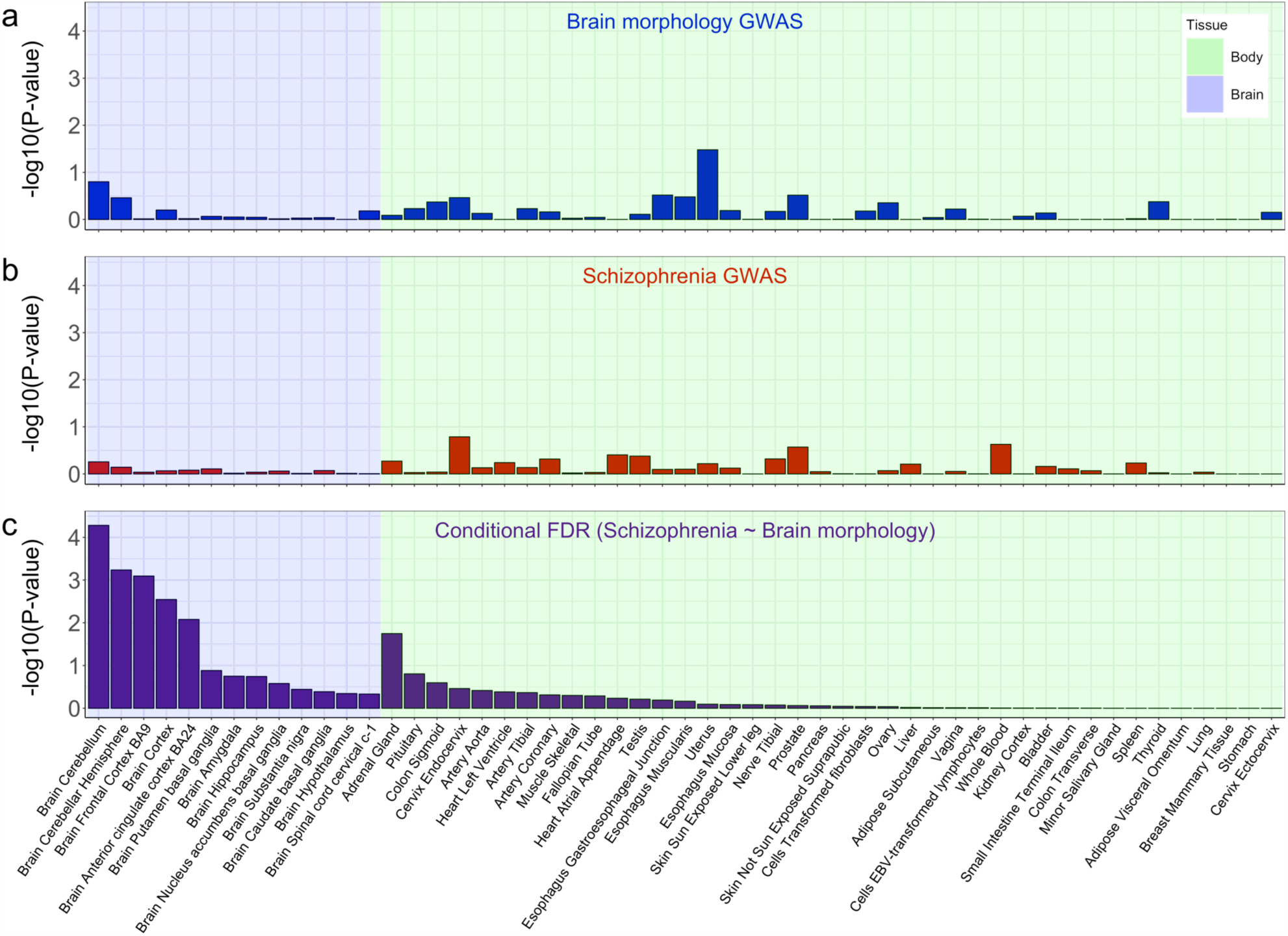
Brain tissue-specific expression of the mapped genes. a) shows the results for the genes mapped through the multivariate brain morphology GWAS, b) the PGC2 schizophrenia GWAS, and c) the pleiotropy-informed conditional FDR approach. The y-axis shows the -log10(p-values) of the hypergeometric tests, testing for enrichment of the identified genes among gene sets with tissue-specific gene expression based on the GTEX database. The x-axis shows each of the 53 tissues, split by brain and body tissues, and sorted within these categories by the significance of the pleiotropy-informed gene set enrichment.

We subsequently created polygenic scores to predict schizophrenia diagnosis for participants of the Thematically Organized Psychosis (TOP) clinical cohort, consisting of 743 individuals with schizophrenia and 1074 healthy individuals. We based the polygenic scores on the PGC2 schizophrenia GWAS summary statistics (excluding the TOP cohort), selecting the most significant lead SNPs identified by either 1) the multivariate brain morphology GWAS, 2) the PGC2 schizophrenia GWAS, or 3) the pleiotropy-informed cFDR analysis. As shown in Figure 3, the pleiotropy-informed polygenic scores were the most predictive, outperforming the PGC2 schizophrenia-based polygenic scores by more than 10% at each SNP inclusion threshold. We replicated these patterns in an independent UKB sample consisting of 576 individuals with a schizophrenia diagnosis and 317,139 individuals without schizophrenia, see Supplementary Figure 5. We based the scores on the number of lead SNPs rather than on significance thresholds to allow for direct comparison between the approaches, at equal numbers of SNPs in each set. However, the pattern remained the same when using common polygenic scoring significance thresholds rather than a set number of lead SNPs, see Supplementary Figure 6. Supplementary Table 3 lists the number of loci overlapping between the approaches at each threshold, and Supplementary Figure 7 shows the distribution of schizophrenia diagnosis odds ratios and standard errors across the SNPs that went into the scores, estimated in the TOP sample.

**Figure 3.**
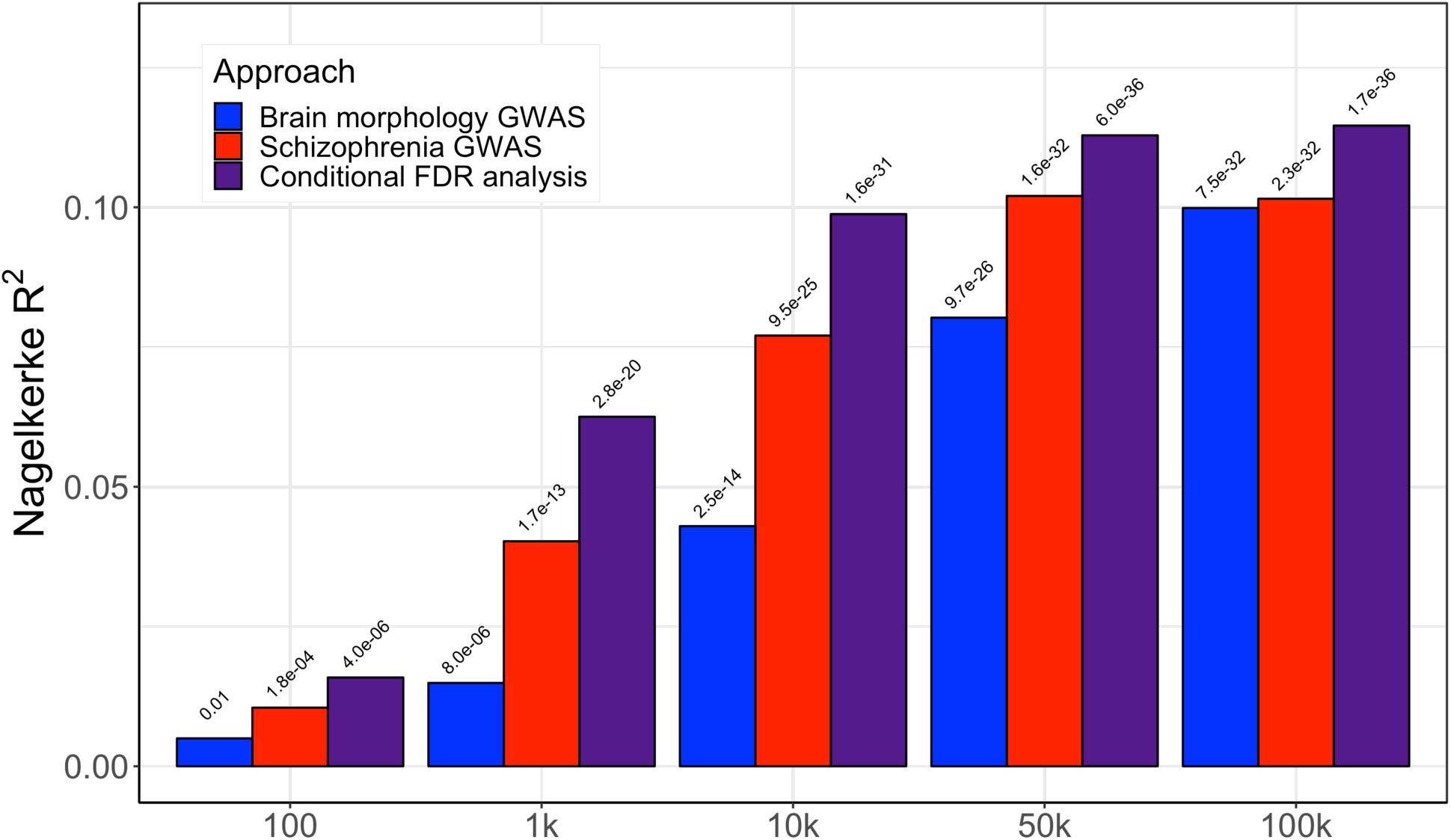
Prediction of schizophrenia diagnosis by polygenic scores, in the TOP sample. The x-axis indicates the number of most significant lead SNPs that went into the polygenic scores, based on the results from the brain morphology GWAS (blue bars), the PGC2 schizophrenia GWAS (in red), or the conditional FDR analysis (in purple). The y-axis indicates Nagelkerke R^2^. The -log10(p-values) of the association between the polygenic scores and schizophrenia diagnosis are listed above the bars.

With the present study, we provide evidence of large genetic overlap between schizophrenia and brain morphology, contrary to a previous report.^10^ In addition to increased sample size, this may result from the application of analysis approaches that are more in line with the complexity of the polygenic architecture of schizophrenia and its overlap with brain measures. In particular, the distributed nature of genetic signal relevant to schizophrenia across the brain and the mixed directions of effects support the use of multivariate approaches and measures of genetic overlap that do not rely on globally consistent directionality of effects.^13,14^ This is exemplified by our high yield in terms of genome-wide significant loci, as well as by our finding that the variants identified were linked to brain tissue-specific gene expression.

We further show, in two independent samples, that prediction of schizophrenia diagnosis improved by conditioning schizophrenia GWAS on association with brain morphology. Possibly, this enrichment approach allows for the reduction of noise in GWAS signal and the identification of more biologically relevant genetic variants, leading to better out-of-sample prediction. This is conceptually similar to a previous report that polygenic scores based on genetic variants specific to placental tissue are more predictive of schizophrenia than those based on non-specific variants.^23^ This may provide a general strategy to improve genetics-based prediction of polygenic brain disorders.

## Data Availability

The data incorporated in this work were gathered from public resources, and the Thematically Organised Psychosis study. The code is available via https://github.com/precimed (GPLv3 license). Correspondence and requests for materials should be addressed to

## Author contributions

D.v.d.M. conceived the study; D.v.d.M., and T.K. pre-processed the data; D.v.d.M. and A.A.S performed all analyses, with conceptual input from T.K., O.F., J.M.S., O.A.A. and L.T.W.; All authors contributed to interpretation of results; D.v.d.M. drafted the manuscript and all authors contributed to and approved the final manuscript.

## Acknowledgements

The authors were funded by the Research Council of Norway (276082, 213837, 223273, 204966/F20, 229129, 249795/F20, 225989, 248778, 249795, 298646, 300767), the South-Eastern Norway Regional Health Authority (2013-123, 2014-097, 2015-073, 2016-064, 2017-004), Stiftelsen Kristian Gerhard Jebsen (SKGJ-Med-008), The European Research Council (ERC) under the European Union’s Horizon 2020 research and innovation programme (ERC Starting Grant, Grant agreement No. 802998) and National Institutes of Health (R01MH100351, R01GM104400). This work was partly performed on the TSD (Tjeneste for Sensitive Data) facilities, owned by the University of Oslo, operated and developed by the TSD service group at the University of Oslo, IT-Department (USIT). Computations were also performed on resources provided by UNINETT Sigma2 -the National Infrastructure for High Performance Computing and Data Storage in Norway.

## Online Methods

### Participants

We made use of data from participants of the UK Biobank (UKB) population cohort, under accession number 27412. The composition, set-up, and data gathering protocols of UKB have been extensively described elsewhere.^15^ After pre-processing, as described below, our sample size for the primary neuroimaging analyses, excluding individuals with schizophrenia, was n=33,735. This sample had a mean age of 64.33 years (SD=7.49), and 52.02% was female. For replication of the polygenic score findings (Supplementary Figure 4) we made use of 576 individuals with a schizophrenia diagnosis (mean age 55.76 years, SD=8.39; 35.60% female) and 317,139 without schizophrenia (mean age 57.43 years, SD=8.04; 54.90% female) that were not part of the neuroimaging sample.

We further used data from the Thematically Organised Psychosis (TOP) clinical cohort. For the polygenic scoring analyses, we had complete genetic data available for 743 individuals with schizophrenia (mean age 32.76 years, SD=13.29; 42.80% female) and 1074 healthy individuals (mean age 32.49 years, SD=10.00; 47.39% female). For the additional analyses of the association of univariate brain measures with diagnosis, we had neuroimaging data available of 457 individuals with schizophrenia (mean age 30.54 years, SD=9.47; 41.79% female) and 998 healthy individuals (mean age 33.33 years, SD=10.26; 45.49% female).

### Genetic data pre-processing

We made use of the UKB v3 imputed data, which has undergone extensive quality control procedures as described by the UKB genetics team.^20^ After converting the BGEN format to PLINK binary format, we additionally carried out standard quality check procedures, including filtering out individuals with more than 10% missingness, SNPs with more than 5% missingness, SNPs with an INFO score below 0.8, and SNPs failing the Hardy-Weinberg equilibrium test at p=1*10^−9^. We further set a minor allele frequency threshold of 0.005, leaving 9,061,072 SNPs.

TOP DNA samples obtained from blood or saliva were sent for genotyping in six separate batches between the years of 2014 and 2017 at deCODE Genetics, Reykjavik, Iceland. The samples were analyzed using Illumina Human OmniExpress12 (HOE12, first four batches), Infinium OmniExpress24 (IOE24, fifth batch) and Infinium Global Screening Array (IGSA, sixth batch) platforms, respectively. Similar but independent procedures were followed for sample batches genotyped on different platforms. We used PLINK to perform pre-imputation quality control, including removal of any samples with possible contamination (heterozygosity more than five standard deviations above the mean) or too low coverage (<80%) to enable its detection and any variants with genotyping rate lower than 95%, Hardy-Weinberg disequilibrium test p-value lower than 10^−4^, high rate of Mendel errors in eventual trios or significant (FDR<0.5) batch effects (in case multiple batches were being processed simultaneously). The quality controlled genotypes were phased using Eagle,^24^ and missing variants were imputed with MaCH^25^ using version 1.1 of the trans-ethnic reference sample assembled by the haplotype reference consortium (HRC). High quality variant sets from the quality control procedure were selected to compute each individual’s genetic principal components and to impute each individual’s sex (X chromosome variants only). Following the quality control and imputation procedure, SNPs with an INFO score lower than 0.8 or minor allele frequency lower than 0.01 were removed. In addition, individual genotypes imputed with less than 75% confidence were set to missing, the remaining ones were converted to best guess hard allelic dosage calls. Finally, individuals were excluded when they had an imputation rate lower than 95%, when there were relatives (up to third-degree) that had a better imputation rate, or when their annotated and imputed sex differed.

### Image acquisition

For the genetic analyses of neuroimaging data, we made use of MRI data from UKB released up to March 2020. T1-weighted scans were collected from four scanning sites throughout the United Kingdom, all on identically configured Siemens Skyra 3T scanners, with 32-channel receive head coils. The UKB core neuroimaging team has published extensive information on the applied scanning protocols and procedures, which we refer to for more details.^16^

For the association of brain morphology with schizophrenia diagnosis, we made use of T1-weighted data from TOP, collected on three different scanners at the Oslo University Hospital, namely a 1.5 T Siemens Magnetom Sonata, a 3.0T General Electric (Signa HDxt), and a 3.0T General Electric (GE750). For details on scanning protocols please see the listed references.^26,27^

### Neuroimaging data pre-processing

The T1-weighted scans were stored locally at the secure computing cluster of the University of Oslo. We applied the standard 舠recon-all -all舡 processing pipeline of Freesurfer v5.3, performing automated surface-based morphometry and subcortical segmentation.^17,18^ From the output, we extracted a total of 175 global, subcortical and cortical morphology measures. Supplementary Table 1 contains all the measures included in the current study. For each of these, we included both the left and right hemisphere measure, if applicable.

For the imaging GWAS of UKB data, we first selected all individuals with White European ancestry, as determined by a combination of self-identification as ‘White British’ and similar genetic ancestry based on genetic principal components, with good quality genetic data. We excluded anyone with an ICD10 schizophrenia diagnosis, as indicated by an ‘F2’ code in UKB data fields 41202 or 41204. We further excluded individuals with bad structural scan quality as indicated by an age and sex-adjusted Euler number (a measure of segmentation quality)^19^ more than three standard deviations lower than the scanner site mean, or with a global brain measure more than five standard deviations from the sample mean. Finally, we removed one of each genetically related pair of individuals, as defined by a threshold of 0.0625 determined by genome-wide complex trait analysis (GCTA).

We followed the same pre-processing procedure for the association of brain measures with schizophrenia diagnosis in the TOP cohort. White European ethnicity was based on self-report. DSM-IV diagnosis of schizophrenia was determined based on the Structured Clinical Interview for DSM-IV Axis I Disorders (SCID), carried out by trained physicians or clinical psychologists. Controls were individuals without brain damage or a lifetime history of a severe psychiatric disorder themselves or first-degree relatives.

For both samples, separately, we regressed out age, sex, scanner site, Euler number, and the first twenty genetic principal components from each brain measure. We further regressed out a global measure specific to each of the feature subsets: eICV for the subcortical volumes, mean thickness for the regional thickness measures, and total surface area for the regional surface area measures. Following this, we applied rank-based inverse normal transformation^28^ to the residuals of each brain measure.

### Genetic analyses

The univariate GWAS on each of the 175 pre-residualized and normalized brain morphology measures were carried out using the standard additive model of linear association in PLINK2^29^. We further carried out multivariate GWAS on all brain morphology measures, using the multivariate omnibus statistical test (MOSTest),^14^ see https://github.com/precimed/mostest.

We made use of the summary statistics from the 2014 PGC2 schizophrenia GWAS^2^ to check for genetic overlap between brain morphology and schizophrenia. We selected a version of the meta-analyzed summary statistics where the TOP sample was left out, to prevent sample overlap in downstream analyses. This version contained 16,052,698 SNPs for 35,099 individuals with schizophrenia and 46,436 controls.

We conducted conditional false discovery rate (cFDR) analysis, conditioning the PGC2 schizophrenia GWAS on the brain morphology GWAS, through the pleioFDR tool using default settings, see https://github.com/precimed/pleiofdr. We set an FDR threshold of 0.01 as whole-genome significance, in accordance with recommendations.

Independent significant SNPs and genomic loci were identified from each of the resulting GWAS summary statistics in accordance with the Functional Mapping and Annotation of GWAS (FUMA) SNP2GENE definition.^22^ We also used the FUMA online platform (https://fuma.ctglab.nl/) to further process the GWAS summary statistics, including the mapping of the lead SNPs to genes, using positional mapping (10kb to gene) and eQTL mapping, based on the eQTL catalogue. As the raw cFDR output is incompatible with FUMA, we first identified lead SNPs according to FUMA definitions by clumping these summary statistics at an FDR threshold of 0.01 (see https://github.com/precimed/python_convert), before uploading to FUMA using the ‘pre-defined lead SNPs’ option.

We further carried out MAGMA-based gene analyses for the GWAS of brain morphology and schizophrenia (not for the cFDR analysis, as this is not valid), through the application of a SNP-wide mean model and use of the 1000 Genomes Phase 3 EUR reference panel.

Hypergeometric tests were applied, through FUMA’s GENE2FUNC function, to calculate the significance of overlap between the three sets of GWAS mapped genes and the sets of differentially expressed genes (DEG) in each of the 53 tissues available in the GTEXv6 database. DEGs are defined as genes with log2 transformed, normalized expression values (Read Per Kilobase per Million, zero-mean) with P-value ≤ 0.05 after Bonferroni correction and absolute log fold change ≥ 0.58 in a given tissue.

For estimates of genetic correlation (rg), we applied cross-trait linkage disequilibrium score regression (LDSR)^30^. For this, each set of summary statistics underwent additional filtering, including the removal of all SNPs in the extended major histocompatibility complex (MHC) region (chr6:25舑35 Mb). To estimate genetic overlap between each brain measure and schizophrenia, regardless of direction of effects, we applied a causal mixture model to the GWAS summary statistics through the bivariate MiXeR tool, see https://github.com/precimed/mixer. We calculated the Dice coefficient as a measure of genetic overlap, based on the ratio of estimated number of causal variants with an effect on both traits to total number of variants involved in either trait.

### Polygenic scoring

We calculated polygenic scores through PRSice v2.^31^ For this, we first clumped each of the three summary statistics (brain morphology, schizophrenia, and the cFDR analysis) through PLINK, using a minor allele frequency threshold of .05, a p-value threshold of 1, an LD threshold of 0.1, and a distance threshold of 10mb. We then ran PRSice, calculating the polygenic scores for the individuals in the TOP sample (and UKB replication sample) based on the PGC2 schizophrenia GWAS summary statistics, using only the most significant lead SNPs from each of the three clumped summary statistics as determined by the indicated PRS threshold. For example, we took the top 10,000 most significant lead SNPs from each of the three pre-clumped summary statistics for the ‘10k’ threshold shown in Figure 3, and every lead SNP with a p-value < .05 for the p=.05 threshold shown in Supplementary Figure 6. Note, we took a maximum threshold of 100,000 SNPs, as this was close to the total number of lead SNPs remaining following clumping, i.e. at p=1 (see Supplementary Figure 6).

We subsequently used logistic regression to calculate the log odds and Nagelkerke R^2^ of the polygenic scores at each threshold, with schizophrenia diagnosis as outcome in the TOP and UKB cohorts separately. We included age, sex and the first twenty genetic principal components as covariates.

### Statistical analyses

All downstream analyses were carried out in R v3.6.1. Graphs were created using *ggplot2*,^32^ and brain maps through pysurfer https://pysurfer.github.io/. Code is available upon request to the corresponding author.

## Supplementary Information

Supplementary Table 1: List of the 175 brain measures included in the analyses

Supplementary Table 2: Overview of number of loci per approach

Supplementary Table 3: Number of PRS lead SNPs overlapping between approaches

Supplementary Table 4: Inference on distribution of test statistics PRS lead SNPs

Supplementary Data 1: List of significant loci from the brain morphology GWAS

Supplementary Data 2: List of significant genes from the brain morphology GWAS

Supplementary Data 3: List of significant loci from the conditional FDR analysis

Supplementary Figure 1: Conditional FDR Manhattan plot

Supplementary Figure 2: Relation genetic overlap brain measures with SZ diagnosis

Supplementary Figure 3: Cortical maps of genetic overlap with SZ diagnosis Supplementary Figure 4: Gene set enrichment in GWAS catalog

Supplementary Figure 5: UKB replication of PRS results Supplementary Figure 6: PRS results using significance thresholds

Supplementary Figure 7: Distribution test statistics of PRS lead SNPs predicting SZ diagnosis

Supplementary Figure 8: Quintile plot PRS lead SNPs predicting SZ diagnosis

### Morphological features included

Supplementary Table 1 lists all brain morphology measures used in the analyses. We included all regional measures outputted by the default FreeSurfer cortical processing streams; these consist of 34 regions per hemisphere, for both hemispheres, for both cortical thickness and surface area, i.e. 136 measures. We further included the most commonly used measures from the subcortical segmentation (’aseg’), namely five global measures and 34 regional subcortical volumes.

**Supplementary Table 1.**
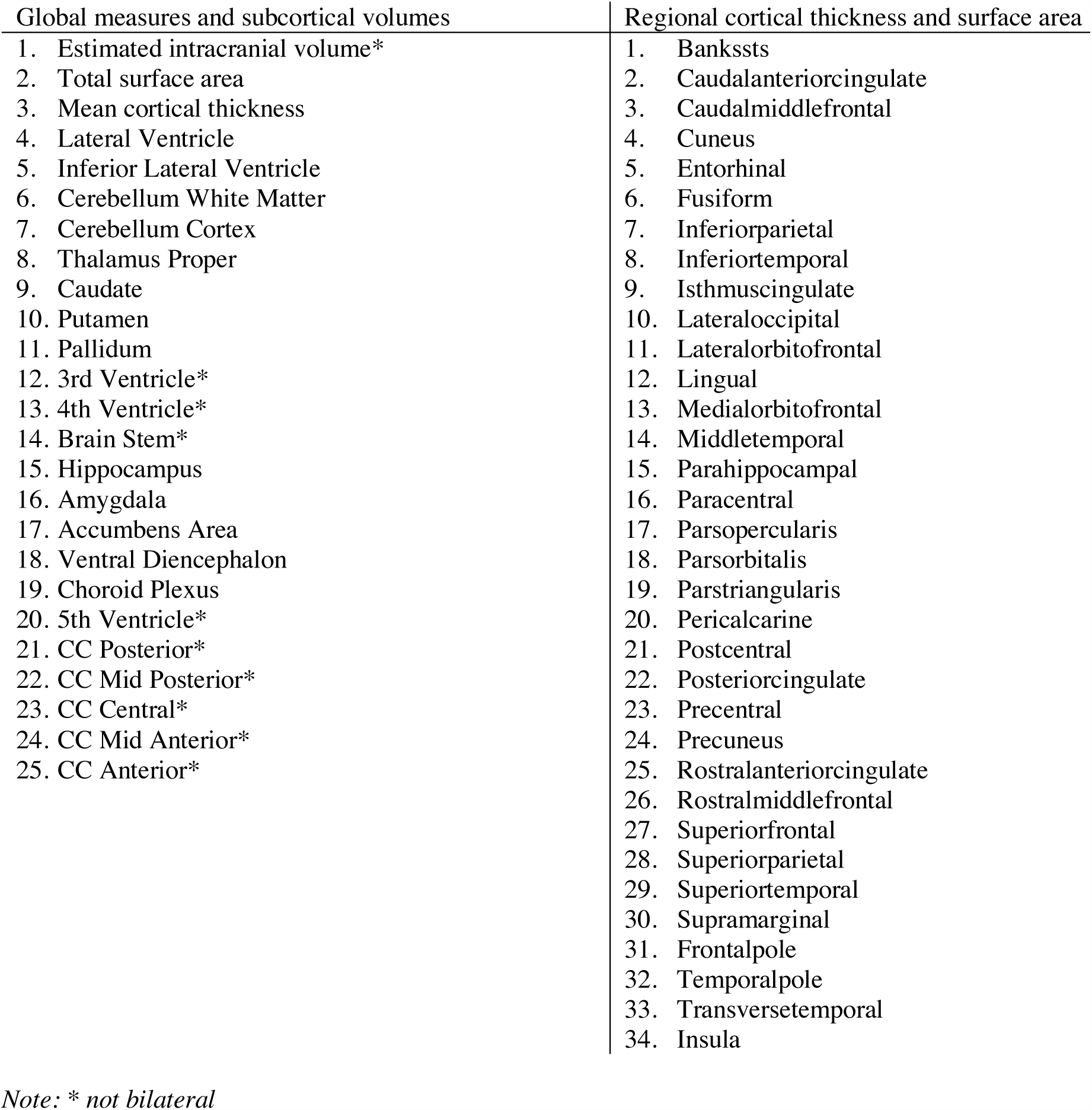
Brain morphology measures included in the study.

### List of discovered loci

Supplementary Table 2 provides an overview of the number of whole-genome significant SNPs, independent SNPs, independent loci, and mapped genes per approach. Supplementary Data 1 contains an overview of each of the individual loci discovered by the multivariate brain morphology GWAS, including information on genomic location, significance, and mapped genes, as outputted by FUMA. This table further lists their p-value as reported by the PGC2 schizophrenia GWAS. Supplementary Data 2 lists the genes found to be significant in the brain morphology GWAS by MAGMA. Supplementary Data 3 contains the information on the loci discovered by the cFDR approach.

**Supplementary Table 2.** Number of whole-genome significant SNPs, independent SNPs, independent loci and mapped genes discovered, per approach.

| Approach | # significant SNPs | # independent SNPs | # independent loci | # genes mapped |
| --- | --- | --- | --- | --- |
| Brain morphology | 99110 | 2722 | 571 | 2245 |
| Schizophrenia | 15800 | 342 | 96 | 458 |
| cFDR | 15422 | 379 | 295 | 836 |

**Supplementary Figure 1.**
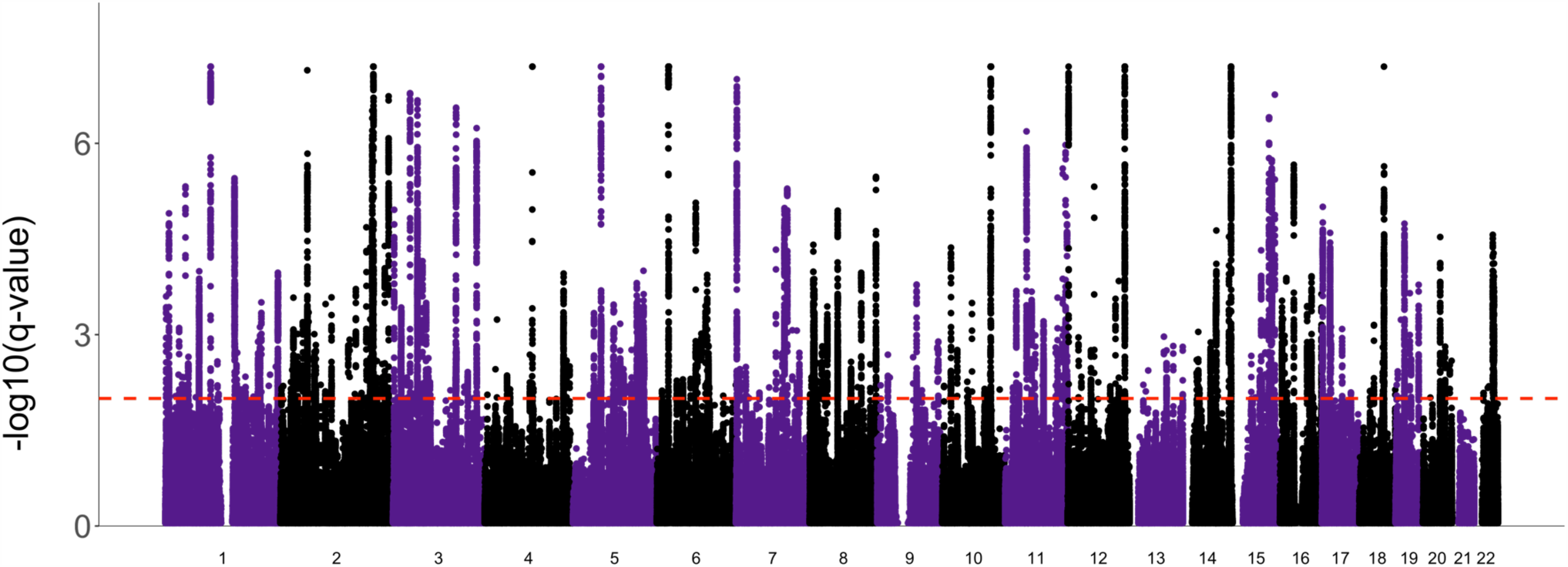
Manhattan plot of the conditional false discovery rate (FDR) analysis, conditioning schizophrenia on brain morphology. The observed -log_10_(q-value) of each SNP is shown on the y-axis, the x-axis shows the relative genomic location grouped by chromosome, and the red dashed lines indicate the whole-genome significance FDR threshold of 0.01.

### Relation between measures of genetic overlap

We estimated the genetic overlap between schizophrenia and each of the univariate brain measures, to complement the multivariate analysis. For this, we took the same PGC2 schizophrenia GWAS summary statistics as used for the primary analyses, excluding the TOP cohort, and each of the 175 univariate brain measure GWAS summary statistics that went into the multivariate analysis. We calculated 1) the genetic correlation through LDSC, 2) the estimated amount of shared causal variants, in the form of a Dice coefficient, through the MiXeR tool, and 3) the number of discovered loci when conditioning the schizophrenia GWAS on the brain measure GWAS, through cFDR. We further ran logistic regression models, predicting schizophrenia diagnosis from each of the brain measures, for participants of the TOP sample with T1-weighted MRI scans (N=457 individuals with schizophrenia, N=998 healthy individuals). Please see the Methods section for more details.

We present the overview of all genetic overlap estimates, and the relation between the brain measures and schizophrenia diagnosis, in Supplementary Figure 2 and 3. Figure 2A-C shows each of the different measurement types on the x-axis, and the brain measures, grouped, on the y-axis. The estimates have been rescaled such that, within each measurement type, the lowest estimate is set to zero and the highest is set to one. For ease of visualization, we show only the left hemisphere. Supplementary Figure 3 provides cortical brain maps, depicting all of the cortical estimates across both hemispheres in, showing that these estimates are similar across both hemispheres. We further calculated the Pearson’s correlation, across all brain measures, between the different measurement types, see Figure 2D. As the Dice coefficients and number of cFDR loci are unidirectional, we first took the absolute of the genetic correlation and diagnosis (log odds) estimates before calculating the correlation with those measurement types.

Across all brain measures, we found that the number of loci discovered through cFDR correlates stronger with schizophrenia diagnosis (Pearson’s R=.45, p=2.9*10^−10^) than the genetic correlation as estimated through linkage disequilibrium score regression (Pearson’s R=0.21, p=0.005).

**Supplementary Figure 2.**
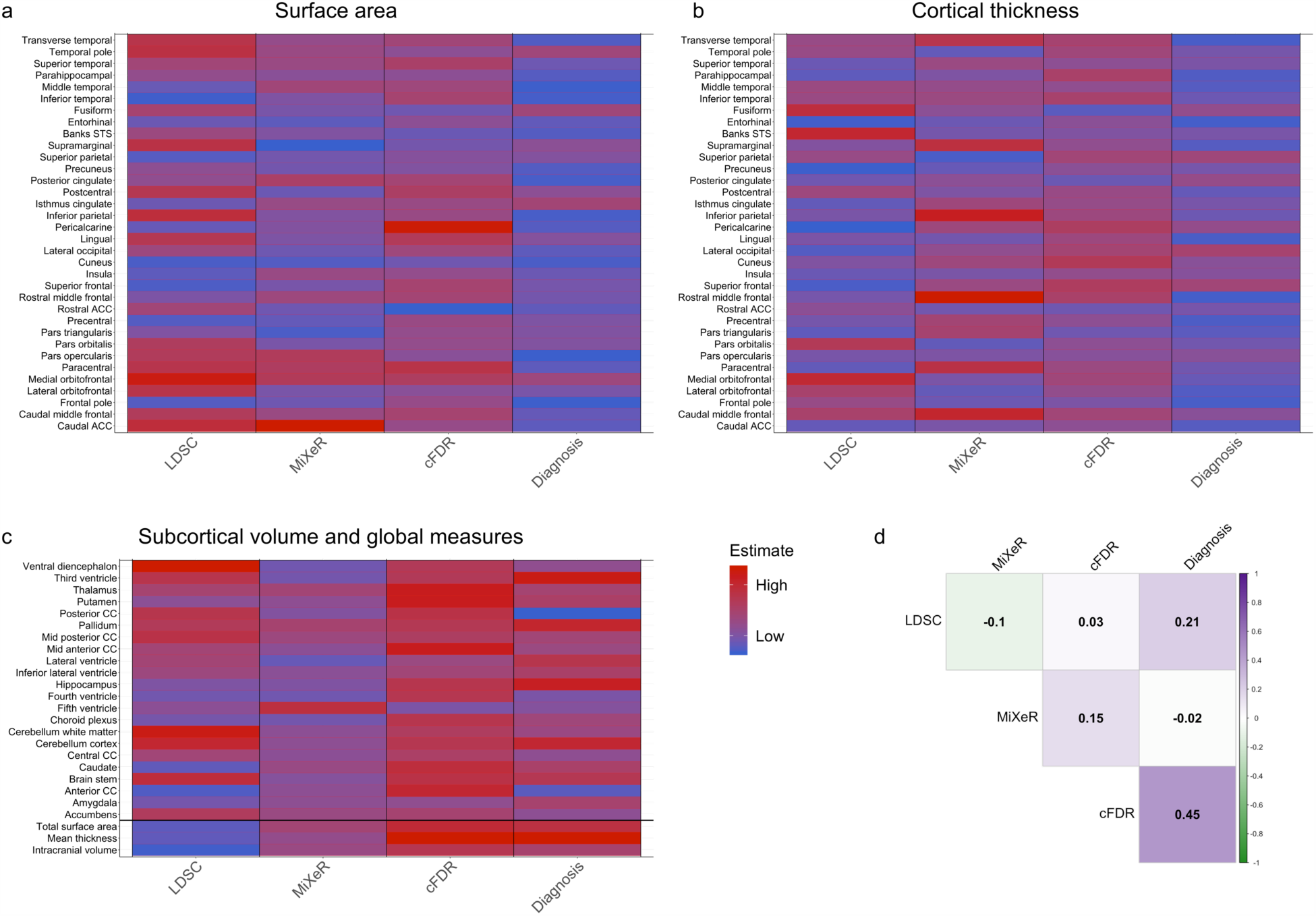
Comparison of genetic overlap between brain measures and schizophrenia with the phenotypic association between brain measures and schizophrenia diagnosis. a-c) show for each brain measure, indicated on the y-axis, the estimates from different approaches to genetic overlap, in addition to the association with schizophrenia diagnosis, indicated on the x-axis. Estimates are scaled from zero to one across all brain measures within each approach to allow for comparison. d) shows the Pearson’s correlation between the estimates from each approach. Note: only left hemispheres are shown for a-c) for ease of visualization. LDSC=linkage disequilibrium score regression; cFDR=conditional false discovery rate; CC=corpus callosum.

**Supplementary Figure 3.**
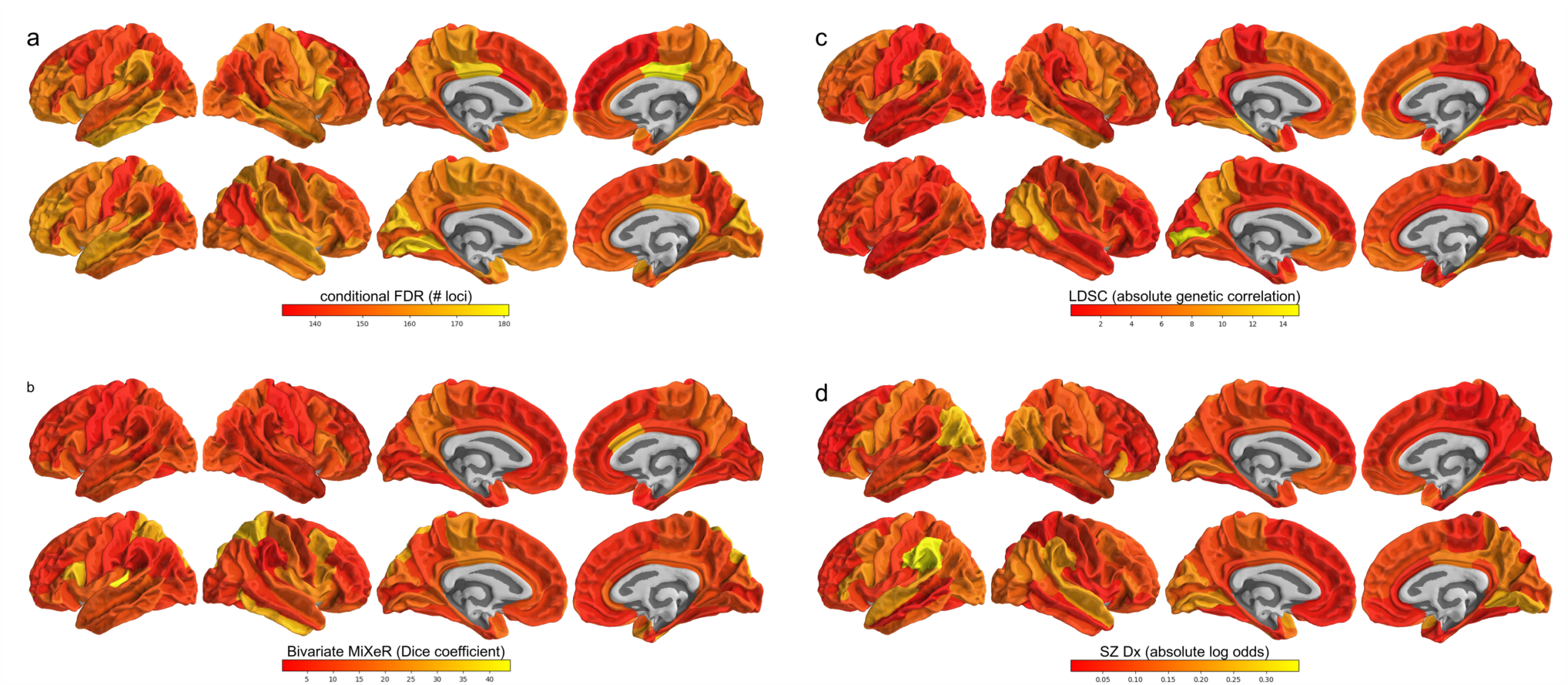
Cortical maps depicting the genetic overlap with schizophrenia per cortical region. a) shows, through color-coding, the number of genome-wide significant loci discovered when conditioning the schizophrenia GWAS on the brain region GWAS, b) shows the Dice coefficient calculated from bivariate MiXeR results, i.e. the proportion of causal genetic variants shared by schizophrenia and each brain region, c) shows the absolute genetic correlation of schizophrenia with each region, as estimated by LD score regression, and d) shows the absolute log odds following from predicting schizophrenia diagnosis by the brain region measures through logistic regression in the TOP sample. The top row of each subplot shows cortical area, and the bottom row shows cortical thickness.

**Supplementary Figure 4.**
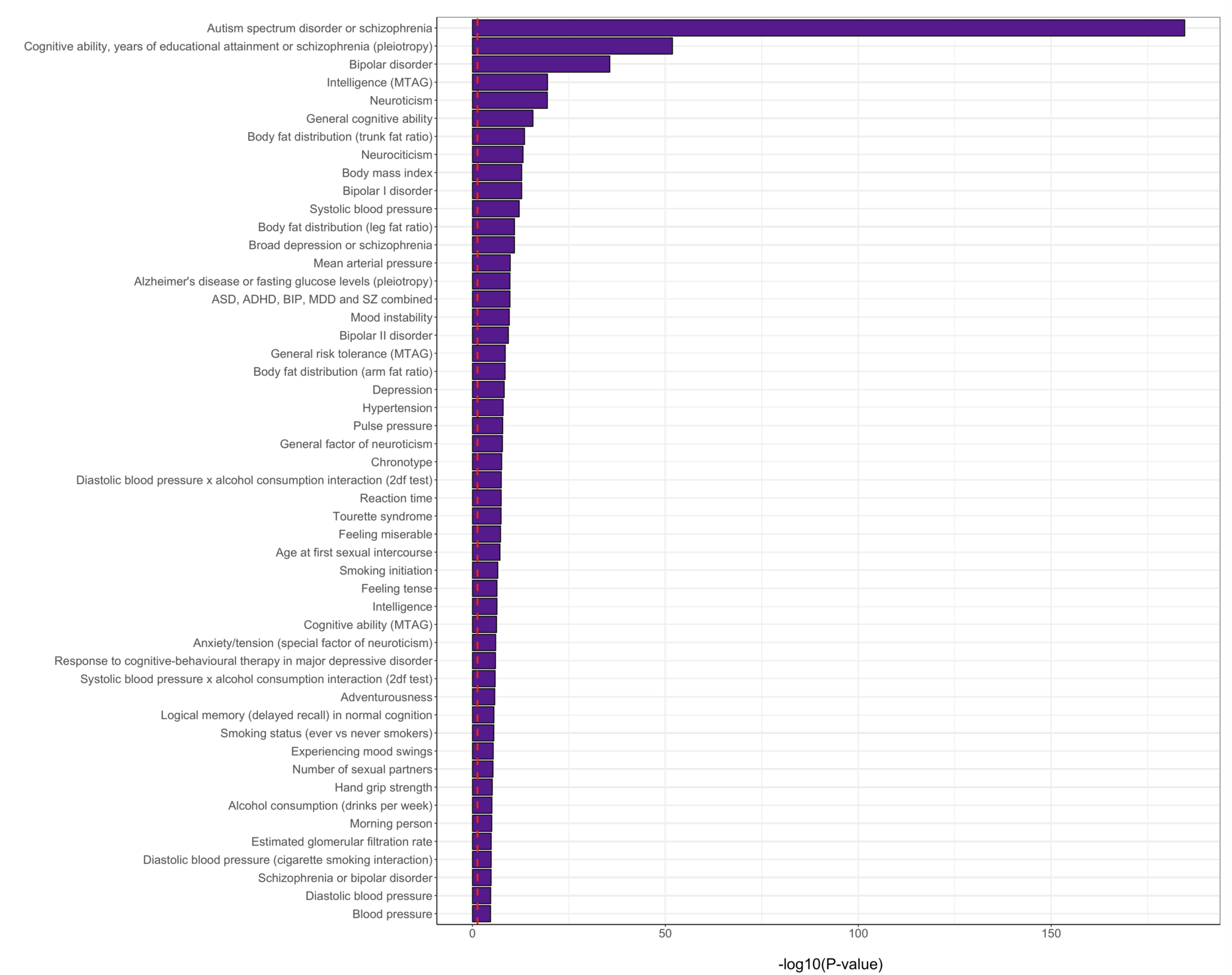
Enrichment of the conditional FDR-mapped genes in GWAS catalog gene sets. On the y-axis are the gene sets identified in previous GWAS as listed in the GWAS catalog, and on the x-axis are the multiple comparisons-corrected -log10(p-values) from hypergeometric tests, testing whether the genes identified through the cFDR analysis are enriched in these sets. Shown are the top 50 most significant gene sets, ordered by their p-value. The red, dashed vertical line indicates the multiple-comparisons corrected significance threshold.

### Replication of polygenic scores in UK Biobank

For replication of the polygenic risk score findings, we used White Europeans in the UK Biobank without neuroimaging data, i.e. those individuals that were not part of the brain morphology GWAS. We then carried out the polygenic scoring pipeline identical as done for the TOP sample. The results pattern was very similar to that found in the TOP sample, as shown in Supplementary Figure 5, with the cFDR analysis-based polygenic scores performing best at each threshold. We do note that the Nagelkerke R^2^ is lower in this sample. This may be due to the UK biobank being a population study rather than a clinical cohort study; individuals with a schizophrenia diagnosis participating in this study may be less severe cases than those included in a clinical study, while the controls are not as stringently selected as in e.g. the TOP study, where anyone was excluded whose first-degree relative had a major psychiatric condition. In addition, the age range of UK biobank may play a role, these are relatively elderly individuals, which may further lessen the comparison with other schizophrenia studies.

**Supplementary Figure 5.**
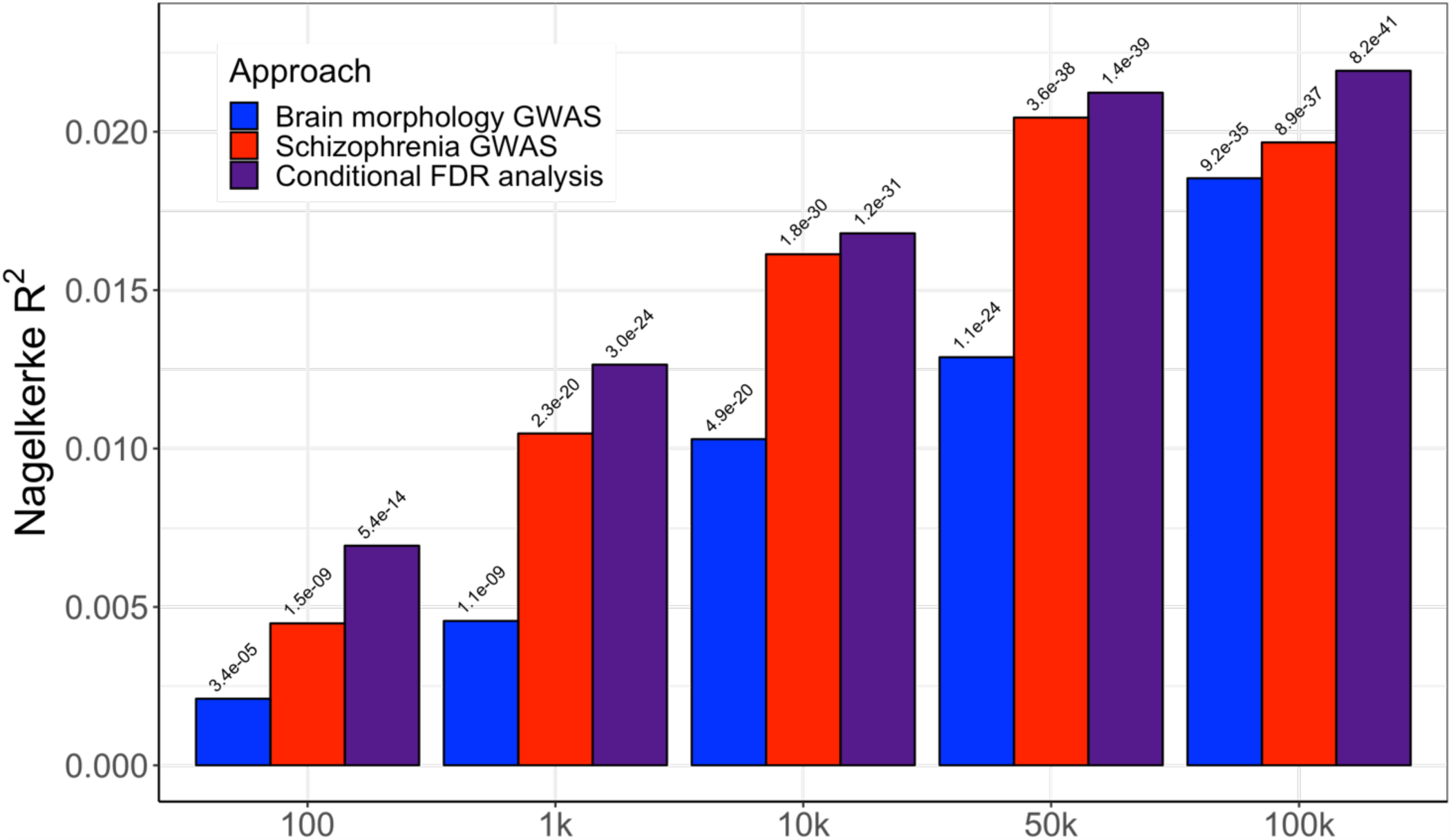
Replication of the polygenic score results in the UK Biobank cohort. The x-axis indicates the number of most significant lead SNPs that went into the polygenic scores, based on the results from the multivariate brain morphology GWAS (blue bars), the PGC2 schizophrenia GWAS (in red), or the conditional FDR analysis (in purple). The y-axis indicates the percent variance explained, Nagelkerke R^2^. The -log10(p-values) of the association between the polygenic scores and schizophrenia diagnosis is listed above each bar.

**Supplementary Figure 6.**
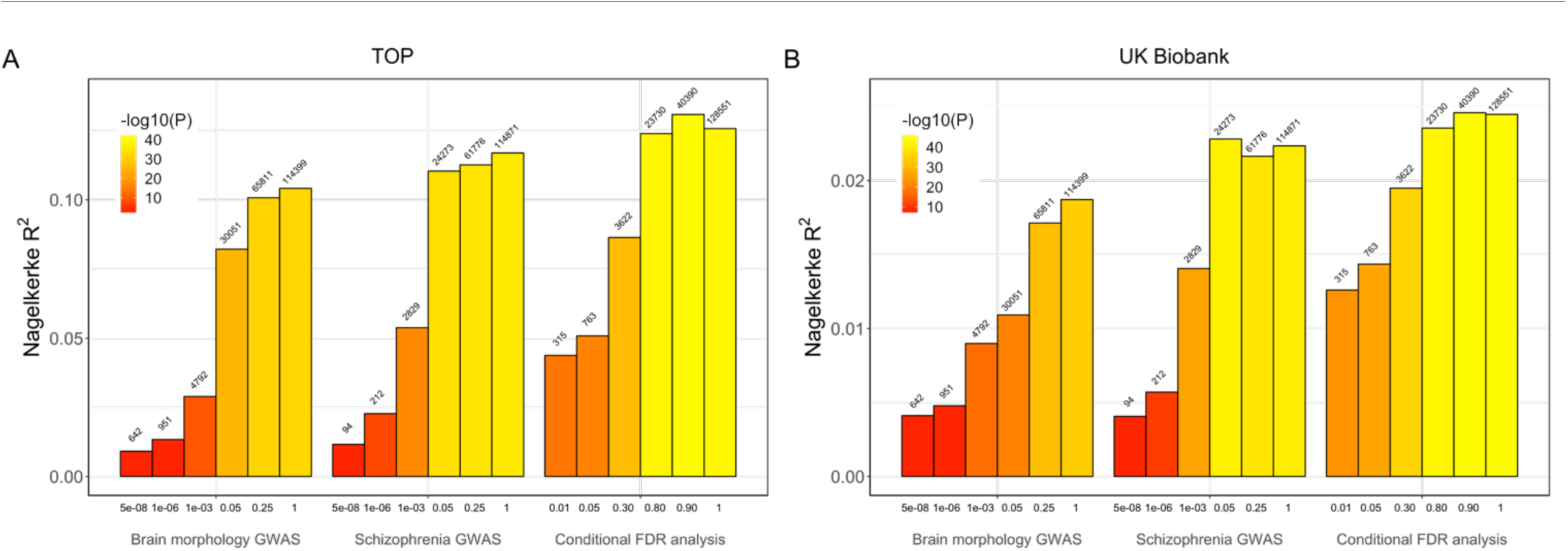
Polygenic scoring results using significance thresholds. A) Results for the TOP sample, B) for the UK Biobank. The x-axis indicates the significance threshold for the SNPs going into the polygenic scores, based on the results from 1) the multivariate brain morphology GWAS, 2) the PGC2 schizophrenia GWAS, or 3) the pleiotropy-informed approach, conditioning the schizophrenia GWAS on the brain morphology GWAS. The y-axis indicates the percent variance explained, Nagelkerke R^2^. The color coding reflects the -log10(p-values) of the association between the polygenic scores and schizophrenia diagnosis. Above each bar is listed how many SNPs went into the score.

### Comparison of sets of loci going into the polygenic scoring analyses

To understand the underlying reason of differences in predictive ability between the three polygenic scoring approaches, we predicted schizophrenia diagnosis from each individual SNP that went into the three polygenic scores at the highest threshold (100k). For this, analogous to the PRS analyses, we used logistic regression with diagnosis as outcome measure, and SNP genotype (number of minor alleles) plus age, sex and twenty genetic components as predictors, in the TOP sample. We then plotted the distributions of absolute log odds and standard errors of the SNP predictors found from these logistic regressions, as shown in Supplementary Figure 7. Supplementary Table 4 shows the results of t-tests checking whether there are significant differences in the means of these three sets of log odds and standard errors. As can be seen, there is no pattern indicating that the pleiotropy-informed cFDR-based polygenic score, which was most predictive, contains on average SNPs with either bigger individual effects or less noise by themselves. This suggests that it is not the individual SNPs that improve the predictive ability, only their joint effect. We speculate that this may be in line with our interpretation of the tissue-specificity of these SNPs; as the cFDR set of SNPs show greater specificity to the brain (Figure 2), they are more likely to act in concert, leading to greater stability across individuals in predicting schizophrenia than sets of SNPs that do not have this biological specificity and therefore have effects scattered across many pathways.

**Supplementary Table 3.**
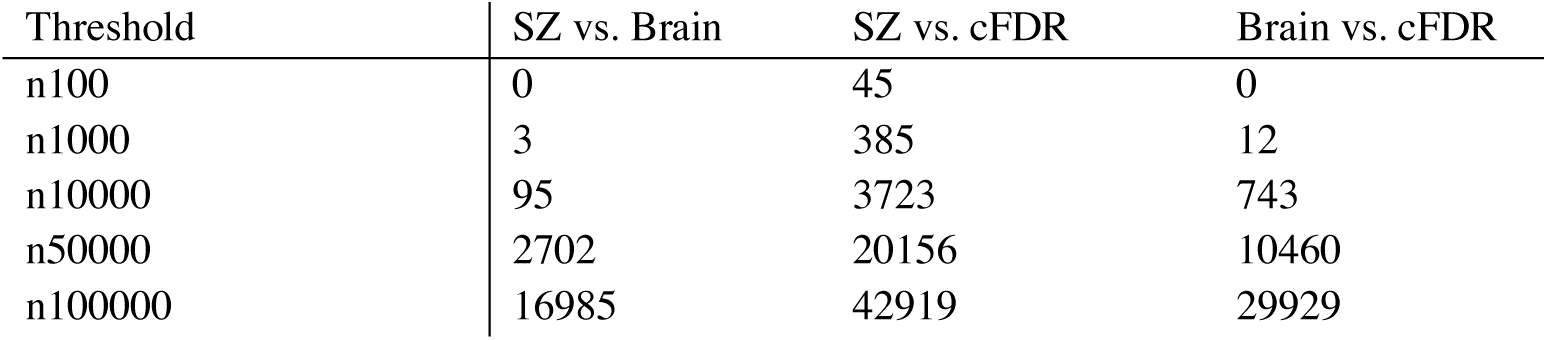
Number of SNPs overlapping between the different polygenic score approaches at each threshold.

| Threshold | SZ vs. Brain | SZ vs. cFDR | Brain vs. cFDR |
| --- | --- | --- | --- |
| n100 | 0 | 45 | 0 |
| n1000 | 3 | 385 | 12 |
| n10000 | 95 | 3723 | 743 |
| n50000 | 2702 | 20156 | 10460 |
| n100000 | 16985 | 42919 | 29929 |

**Supplementary Figure 7.**
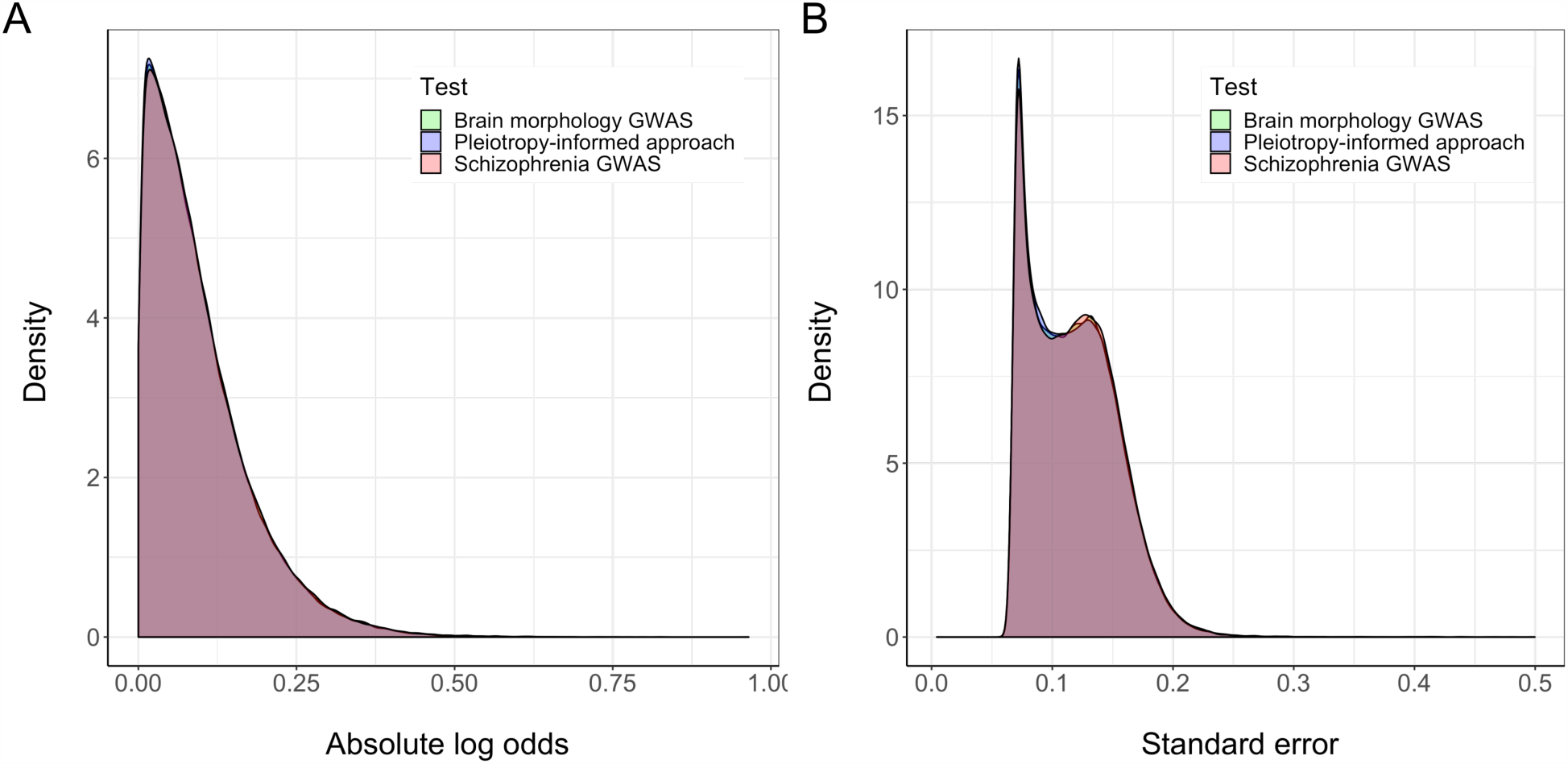
Distribution of test statistics from predicting schizophrenia by the SNPs going into the polygenic scores for the three different approaches. A) The distribution of absolute log odds, B) the distribution of standard errors. These are based on the SNPs that went into the 100k-thresholded PRS.

**Supplementary Table 4.** Results from t-tests on the distributions of log odds and standard errors obtained from logistic regressions predicting schizophrenia diagnosis from each of the 100,000 SNPs that went into the polygenic scores of the three different approaches. The ‘Log odds’ and ‘Standard error’ columns indicates the mean difference between the two approaches specified in the first column, and the p-value columns indicate the associated p-values from the t-test.

| | Log odds | $P_{\log \text{ odds}}$ | Standard error | $P_{SE}$ |
| --- | --- | --- | --- | --- |
| SZ - Brain | 0.001 | 0.11 | 0.017 | 0.14 |
| SZ - cFDR | 0.002 | 4.7e-03 | 0.017 | 0.12 |
| Brain - cFDR | 0.001 | 0.12 | 0.001 | 0.93 |

**Supplementary Figure 8.**
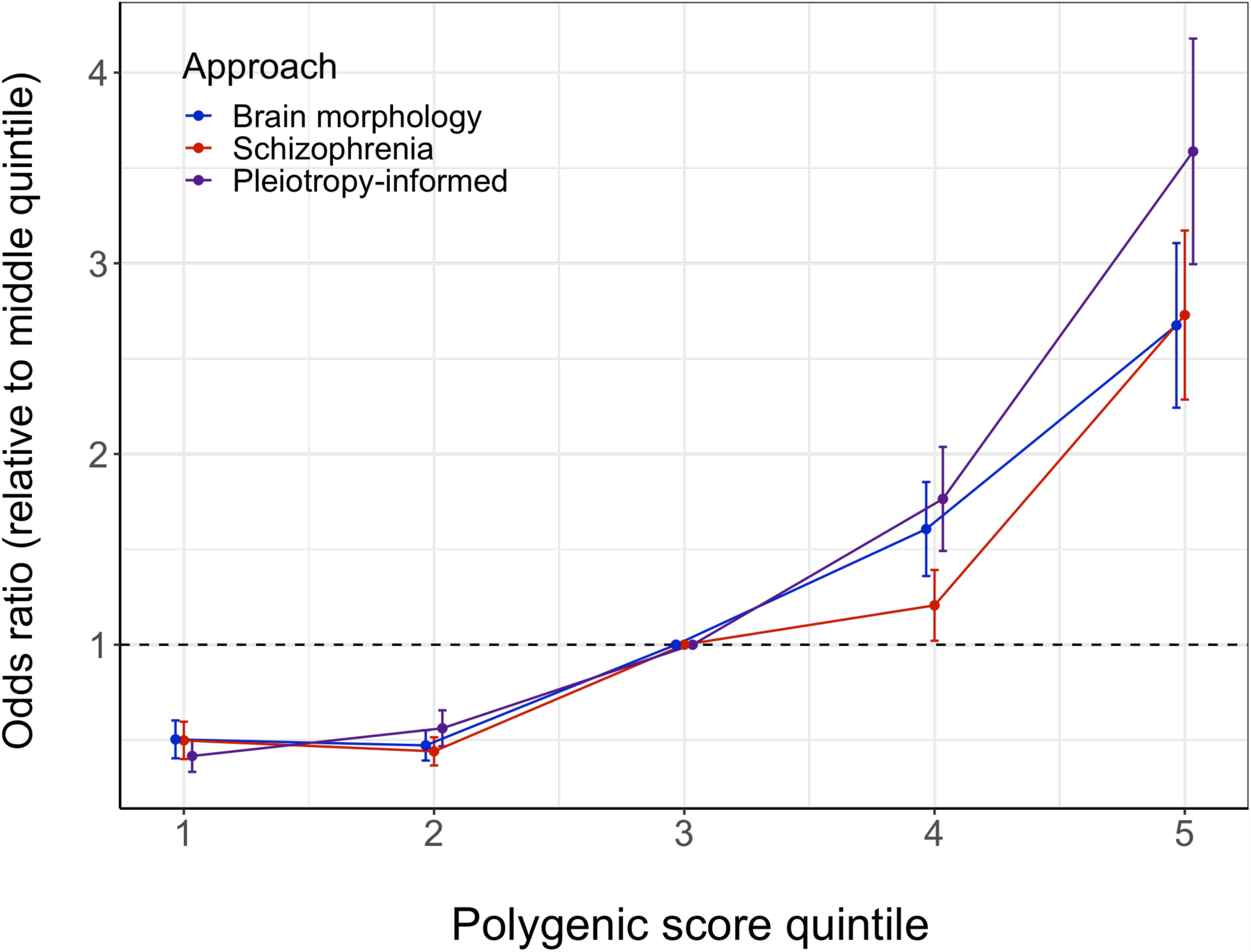
Plot showing the schizophrenia odds ratios of individuals binned into five groups based on their polygenic scores, relative to the middle group. The colors indicate the approach that the polygenic risk score is based on; the vertical lines around the dots indicate the standard errors; the black dashed horizontal line demarcates an odds ratio of 1.

**Supplementary Table 5.** R^2^ on the liability scale for case-control ascertained studies, for the polygenic risk scores. For the main analyses, we presented the Nagelkerke R^2^ of the scores, as most commonly used in polygenic scoring studies, including the PGC2 schizophrenia GWAS, thereby allowing for easy comparison with these studies. However, Nagelkerke R^2^ is based on the disease scale rather than a liability scale and, in addition, it is not invariant to ascertainment. Here, we present an R^2^ on the liability scale that takes into account the schizophrenia case-control ratio in the TOP sample (p=.41) and in the general population (k=.01). For more information, please see Lee et al. 2012.^33^

| # SNPs | Brain morphology GWAS | Schizophrenia GWAS | cFDR |
| --- | --- | --- | --- |
| 100 | 0.002 | 0.004 | 0.007 |
| 1k | 0.006 | 0.017 | 0.027 |
| 10k | 0.019 | 0.034 | 0.044 |
| 50k | 0.036 | 0.046 | 0.051 |
| 100k | 0.045 | 0.046 | 0.052 |

## References

1. Smeland, O., Frei, O., Dale, A. & Andreassen, O. A. The polygenic architecture of schizophrenia – rethinking pathogenesis and nosology. Nat. Rev. Neurol.

2. Consortium, S. W. G. of the P. G. Biological insights from 108 schizophrenia-associated genetic loci. Nature 511, 421 (2014).

3. Holland, D. et al. Beyond SNP heritability: Polygenicity and discoverability of phenotypes estimated with a univariate Gaussian mixture model. PLOS Genet. 16, e1008612 (2020).

4. van Erp, T. G. M. et al. Cortical Brain Abnormalities in 4474 Individuals With Schizophrenia and 5098 Control Subjects via the Enhancing Neuro Imaging Genetics Through Meta Analysis (ENIGMA) Consortium. Biol. Psychiatry 84, 644– 654 (2018).

5. van Erp, T. G. M. et al. Subcortical brain volume abnormalities in 2028 individuals with schizophrenia and 2540 healthy controls via the ENIGMA consortium. Mol Psychiatry 21, 547–553 (2016).

6. Moberget, T. et al. Cerebellar volume and cerebellocerebral structural covariance in schizophrenia: a multisite mega- analysis of 983 patients and 1349 healthy controls. Mol. Psychiatry 23, 1512–1520 (2018).

7. Alnæs, D. et al. Brain Heterogeneity in Schizophrenia and Its Association With Polygenic RiskBrain Heterogeneity in Schizophrenia and Polygenic RiskBrain Heterogeneity in Schizophrenia and Polygenic Risk. (2019). doi:10.1001/jamapsychiatry.2019.0257

8. Wolfers, T. et al. Mapping the heterogeneous phenotype of schizophrenia and bipolar disorder using normative models. JAMA psychiatry 75, 1146–1155 (2018).

9. Andreassen, O. A., Thompson, W. K. & Dale, A. M. Boosting the power of schizophrenia genetics by leveraging new statistical tools. Schizophr. Bull. 40, 13–17 (2013).

10. Franke, B. et al. Genetic influences on schizophrenia and subcortical brain volumes: large-scale proof of concept. Nat Neurosci 19, 420–431 (2016).

11. Ohi, K. et al. Genetic correlations between subcortical brain volumes and psychiatric disorders. Br. J. Psychiatry 216, 280–283 (2020).

12. Frei, O. et al. Bivariate causal mixture model quantifies polygenic overlap between complex traits beyond genetic correlation. Nat. Commun. 10, 2417 (2019).

13. Smeland, O. B. et al. Discovery of shared genomic loci using the conditional false discovery rate approach. Hum. Genet. 139, 85–94 (2020).

14. van der Meer, D. et al. Understanding the genetic determinants of the brain with MOSTest. Nat. Commun. 11, 3512 (2020).

15. Sudlow, C. et al. UK biobank: an open access resource for identifying the causes of a wide range of complex diseases of middle and old age. PLoS Med. 12, e1001779 (2015).

16. Miller, K. L. et al. Multimodal population brain imaging in the UK Biobank prospective epidemiological study. Nat. Neurosci. 19, 1523–1536 (2016).

17. Desikan, R. S. et al. An automated labeling system for subdividing the human cerebral cortex on MRI scans into gyral based regions of interest. Neuroimage 31, 968–980 (2006).

18. Fischl, B. et al. Whole brain segmentation: automated labeling of neuroanatomical structures in the human brain. Neuron 33, 341–355 (2002).

19. Rosen, A. F. G. et al. Quantitative assessment of structural image quality. Neuroimage 169, 407–418 (2018).

20. Bycroft, C. et al. The UK Biobank resource with deep phenotyping and genomic data. Nature 562, 203–209 (2018).

21. de Leeuw, C. A., Mooij, J. M., Heskes, T. & Posthuma, D. MAGMA: generalized gene-set analysis of GWAS data. PLoS Comput. Biol. 11, e1004219 (2015).

22. Watanabe, K., Taskesen, E., Bochoven, A. & Posthuma, D. Functional mapping and annotation of genetic associations with FUMA. Nat. Commun. 8, 1826 (2017).

23. Ursini, G. et al. Convergence of placenta biology and genetic risk for schizophrenia. Nat. Med. 24, 792–801 (2018).

24. Loh, P.-R. et al. Reference-based phasing using the Haplotype Reference Consortium panel. Nat. Genet. 48, 1443 (2016).

25. Li, Y., Willer, C. J., Ding, J., Scheet, P. & Abecasis, G. R. MaCH: using sequence and genotype data to estimate haplotypes and unobserved genotypes. Genet. Epidemiol. 34, 816–834 (2010).

26. Brandt, C. L. et al. Cognitive effort and schizophrenia modulate large-scale functional brain connectivity. Schizophr. Bull. 41, 1360–1369 (2015).

27. Kaufmann, T. et al. Disintegration of Sensorimotor Brain Networks in Schizophrenia. Schizophr. Bull. 41, 1326–1335 (2015).

28. Beasley, T. M., Erickson, S. & Allison, D. B. Rank-based inverse normal transformations are increasingly used, but are they merited? Behav. Genet. 39, 580–595 (2009).

29. Chang, C. C. et al. Second-generation PLINK: rising to the challenge of larger and richer datasets. Gigascience 4, 7 (2015).

30. Bulik-Sullivan, B. et al. An atlas of genetic correlations across human diseases and traits. Nat. Genet. 47, 1236 (2015).

31. Choi, S. W. & O’Reilly, P. F. PRSice-2: Polygenic Risk Score software for biobank-scale data. Gigascience 8, (2019).

32. Wickam, H. ggplot2: elegant graphics for data analysis. New York Springer. http//:had.co.nz/ggplot2/book. Accessed August 5, 2014 (2009).

33. Lee, S. H., Goddard, M. E., Wray, N. R. & Visscher, P. M. A Better Coefficient of Determination for Genetic Profile Analysis. Genet. Epidemiol. 36, 214–224 (2012)

